# *Mycobacterium tuberculosis*-specific T cell responses are impaired during late pregnancy with elevated biomarkers of tuberculosis risk postpartum

**DOI:** 10.1101/2021.06.11.21258789

**Authors:** Aparajita Saha, Jaclyn Escuduero, Troy Layouni, Barbra Richardson, Sharon Hou, Nelly Mugo, Andrew Mujugira, Connie Celum, Jared M. Baeten, Jairam Lingappa, Grace C. John-Stewart, Sylvia M. LaCourse, Javeed A. Shah

**Author notes:** Corresponding author: Javeed A. Shah. Both authors contributed equally (Sylvia M. LaCourse, Javeed A. Shah). Author Contributions: AS: contributed to the design, data acquisition, analysis, and manuscript preparation. JE: data analysis, manuscript preparation, sample collection and transport, TL: statistical analysis, software, BR: data analysis, statistical support, manuscript editing; SH/HH: sample transport, sample storage, database management, NM/AM: sample collection/storage, participant enrollment, CC/JB: cohort enrollment, funding, sample storage, manuscript editing, JL: cohort enrollment, sample storage, funding, manuscript editing, GJS: experimental design, data analysis, funding, manuscript editing, SL: experimental design, data analysis, grant funding, manuscript preparation and editing; JAS: experimental design, data analysis, grant funding, manuscript preparation and editing. Descriptor: 11.4 Mycobacterial Disease: Host Defenses. This title has an online data supplement which is accessible from this issue’s table of content online at www.atsjournals.org.

## Abstract

**Rationale:** Pregnancy is a risk factor for progression from latent tuberculosis infection (LTBI) to symptomatic tuberculosis (TB). However, how dynamic immunologic changes in pregnancy influence immune responses to *M. tuberculosis* (Mtb) is unknown.

**Objectives:** We performed a detailed characterization of Mtb-specific T cell responses of women at high risk for Mtb infection, leveraging a biorepository of longitudinally samples collected before, during, and after pregnancy in high HIV/TB burden settings.

**Methods:** We used specimens collected from women who became pregnant while enrolled in a randomized controlled trial of pre-exposure prophylaxis for HIV prevention. We measured Mtb-specific cytokines, CCR7 and CD45RA memory markers, and overall CD4+ and CD8+ T cell activation from 49 women using COMPASS, a Bayesian statistical method for evaluating overall antigen-specific T cell responses measured by flow cytometry.

**Measurements and Main Results:** 22 LTBI+ women, defined by flow cytometry, demonstrated significantly diminished Mtb-specific CD4+ cytokine responses in the third trimester (COMPASS score (PFS) 0.07) compared before (PFS 0.15), during (PFS 0.13 and 0.16), and after pregnancy (PFS 0.14; p = 0.0084, Kruskal-Wallis test). Paradoxically, Mtb-specific CD8+ cytokine responses and nonspecifically activated CD38+HLA-DR+CD4+ T cells increased during late pregnancy. Nonspecific T cell activation, a previously validated biomarker for progression from LTBI to TB disease, was increased in LTBI+ women postpartum, compared with LTBI-women.

**Conclusions:** Pregnancy-related functional T cell changes were most pronounced during late pregnancy. Mtb-specific T cell changes during pregnancy and postpartum, increases in immune activation may contribute to increased risk for TB progression in the postpartum period.

## Introduction

Tuberculosis (TB) disease in pregnancy and postpartum is associated with poor maternal and infant outcomes (1, 2). Recent large cohort studies demonstrate that pregnancy is associated with increased rates of tuberculosis disease (TB) up to six months postpartum (3, 4). However, the immune mechanisms underpinning this observation are not clear. Successful pregnancy requires dynamic alteration of the local immune environment (5-7). These adaptations to fetal development are accompanied by cell-specific systemic immune alterations (8). Pathogen-specific responses may be influenced by these systemic immune changes, which can alter disease risk (9-11). Understanding pathogen-specific changes to the immune response during pregnancy may improve efforts to prevent and control disease.

T cell responses are a critical aspect of Mtb host defense. Multiple lines of evidence, including studies in mouse and nonhuman primate studies of CD4+ T cell knockout or depletion, human genetic studies, and observational studies during HIV infection demonstrate that CD4+ T cells are essential for Mtb control (12-16). Prior data suggests that IFNγ responses in CD4+ T cells are essential for Mtb protection (17). The frequency of CD4+ T cells simultaneously producing IL-2, TNF, and IFNγ (e.g., that are polyfunctional) is correlated with Mtb protection (18). Other T cell phenotypes, such as central memory cells (Tcm) and CD8+ T cells, may contribute to Mtb host defense (19-22). Conversely, increased total numbers of nonspecifically activated HLA-DR+CD4+ T cells are a correlate of TB progression (23, 24). Since studies of pregnancy commonly begin enrollment during the first or second trimester of pregnancy, they are often unable to capture immune variation prior to and earlier in pregnancy. We leveraged a unique biorepository of samples and data collected longitudinally before, during, and after pregnancy from women in sub-Saharan Africa enrolled in a randomized controlled trial of pre-exposure prophylaxis for HIV prevention to evaluate the effects of various stages of pregnancy on Mtb-specific T cell responses.

## Methods

### Study Population

The protocol for this study was approved by the University of Washington Human Subjects Review Committee and ethics review committee at each study site. All participants provided written informed consent in English or their native language. The Partners PrEP Study was a randomized clinical trial of antiretroviral pre-exposure prophylaxis (PrEP) in HIV serodiscordant couples (25). 4758 total participants were enrolled from 9 sites in Kenya and Uganda between 2007 and 2012. Partners with HIV and CD4 cell count ≥250, without history of AIDS-defining diagnosis or current use of antiretroviral therapy (ART) were eligible. Participants without HIV underwent monthly HIV testing from the parent trial and had urine pregnancy testing at enrollment and monthly thereafter; women living with HIV (WLHIV) had urine collected quarterly. WLHIV not already on ART who became pregnant were referred for prevention of maternal to child transmission services. Participants were eligible for this analysis if they had at least two samples available within 6 months prior to and during incident pregnancy. If available, additional samples were tested from within 6 months postpartum. (**Figure 1A**). For participants with only two timepoints available (before and during pregnancy), paired testing was done only on participants with pregnancy samples from ≥ 20 weeks gestation after interim evaluation of data from participants with three time points samples. WLHIV received antiretrovirals for prevention of maternal to child transmission of HIV, but not for treatment based on the guidelines at the time.

**Figure 1.**
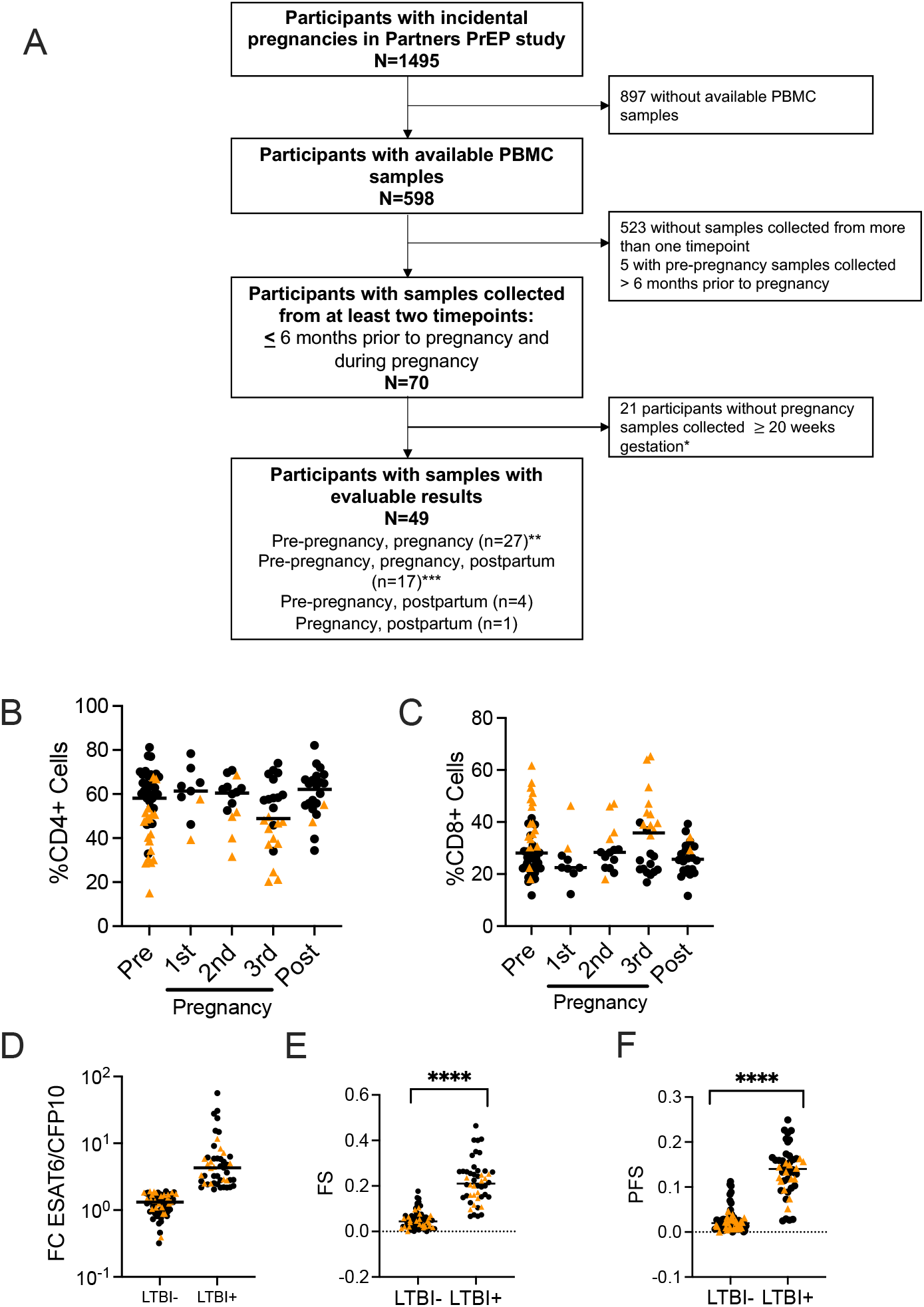
Baseline Immune Characteristics Are Stable Across Pregnancy. A) Flowchart of stored sample selection of participants with incidental pregnancies in the Partners PrEP study. B) Proportion of CD4+ T cells of all CD3+ cells, by pregnancy, trimester, and HIV status. Here and throughout, Samples indicated with *black circles* are from HIV- donors, while *gold triangles* are from WLHIV here and throughout this study. C) Proportion of CD8+ T cells, stratified by pregnancy, trimester, and HIV status. Pre-pregnancy sample N = 49; 1^st^ trimester N = 9, 2^nd^ trimester N = 14; 3^rd^ trimester N = 24; postpartum N = 22. *Black circles* indicate HIV- study participants, while *gold tria*ngles indicate WLHIV here and throughout this paper. Bars indicate median values. Statistical difference between CD4 and CD8+ T cells was measured by Kruskal-Wallis test with sub-analysis comparing column medians. D) Fold change of the proportion of CD4+ T cells expressing IFNγ after incubation with ESAT-6 and CFP-10 pooled peptides, compared to vehicle control. E) COMPASS functional score (FS), stratified by LTBI status. F) COMPASS polyfunctional score (PFS), stratified by LTBI status.

### Statistical Analysis

We used the R package COMPASS to analyze the overall cytokine response of CD4+ and CD8+ T cells and to determine cytokine-producing subsets. COMPASS uses a Bayesian hierarchical framework to model all observed cell subsets and select those most likely to have antigen-specific responses (26, 27). COMPASS provided a functional score (FS), which is the proportion of antigen specific subsets detected among all possible ones, and polyfunctional score (PFS), which is similar, but weighs the different subsets to favor subsets with higher degrees of functionality. Our primary outcome of interest was the COMPASS PFS from CD4+ and CD8+ T cells of LTBI+ women with preexisting T cell responses against Mtb, restimulated with pooled peptides from the Mtb-specific antigens ESAT-6 and CFP10. Differences in median values were compared among timepoints of interest using Kruskal-Wallis tests as the primary analysis, followed by Dunn’s test to compare individual groups. Samples from women with and without HIV were measured together except where indicated.

## Results

### Cohort characteristics and timing of sample collection

We identified 70 participants with incident pregnancy. 49 of these met criteria for this study, with paired samples collected during pregnancy and another, collected less than 6 months prior to the pregnancy. 17 participants had samples collected from 3 timepoints (pre-, during-, and post-pregnancy). 32 participants had samples collected before and during pregnancy (**Figure 1A**). A total of 117 samples with evaluable results were analyzed; 48 pre-pregnancy (median 18.7 weeks [IQR 12.4-22.2), 47 during pregnancy, (median 26.9 weeks gestation [IQR 12.7-30.9]); 22 post-partum (median 23.1 weeks [IQR 18.4-39.6]). The median age of study participants was 26.4 years (IQR 23.2-31.1); 36.7% (18/49) were WLHIV with median CD4 766 cells/mm^3^ [IQR 507-1070]) (**Table 1**).

**Table 1.**
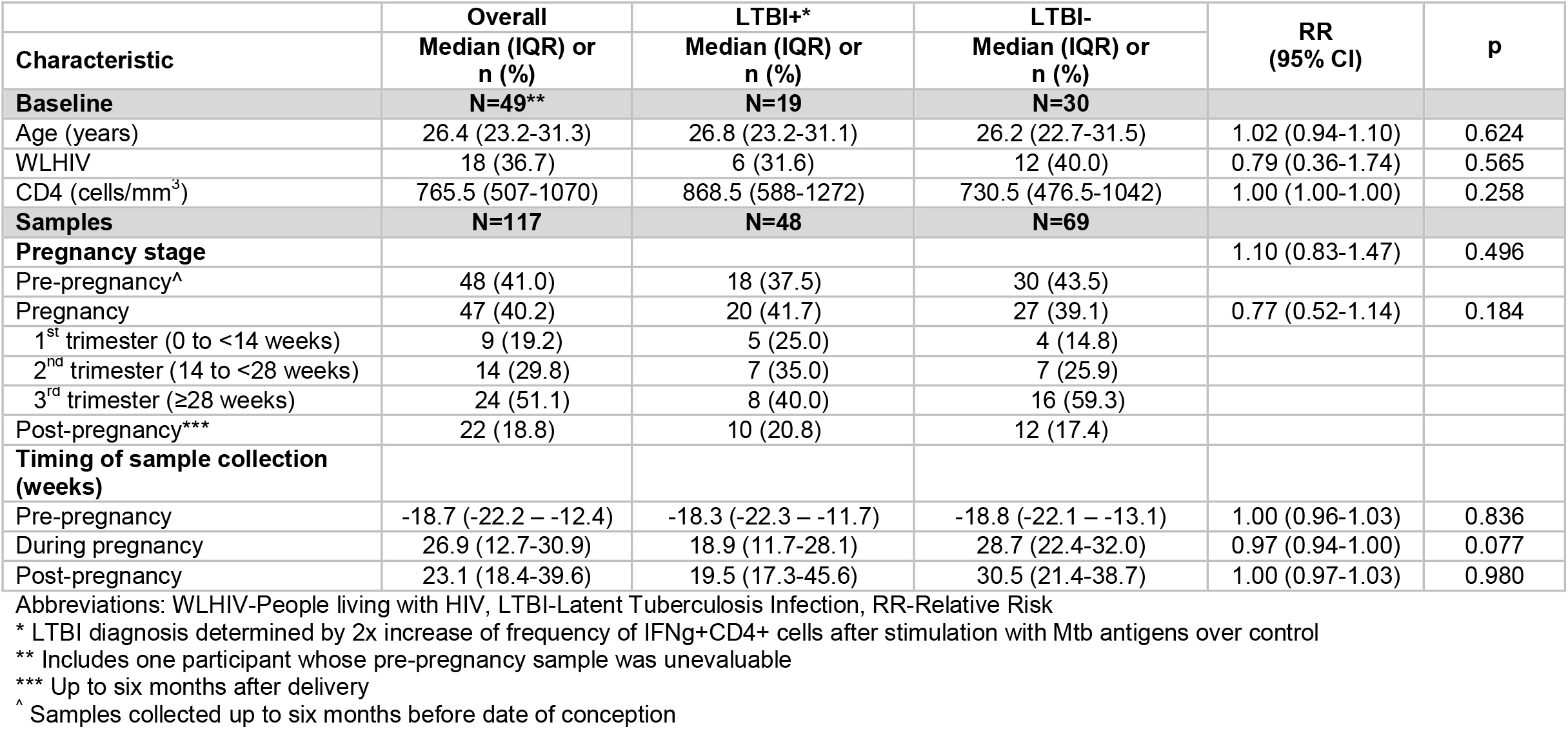
Baseline characteristics of participants and timing of samples with evaluable results.

### Identification of LTBI+ participants using flow cytometry

We identified individuals with LTBI using their first evaluable sample using a flow-based assay as TST or IGRA were not performed during the original trial. We defined individuals as LTBI+ if their proportion of the frequency of IFNγ+CD4+ T cells doubled after ESAT-6 and CFP-10 pooled peptide stimulation compared to DMSO. This measurement correlates well with commercial IGRA tests (28-31). 22/49 (45%) individuals were LTBI+ by this measure (**Figure 1B**). We found that increasing the threshold for positivity to three-fold above background reduced the proportion of LTBI+ individuals slightly, from 22 (44%) to 19 (41%). These observations are similar in frequency to other studies of women of childbearing age in regions with high Mtb burden (32). We used the initially identified more inclusive cutoff to ensure we identified all individuals with pre-existing immune responses to Mtb. We identified three individuals who were LTBI-at baseline but LTBI+ by their pregnancy visit. In these situations, we included these individuals as LTBI+ from their first positive result onward.

### CD4+ and CD8+ T cell frequency is consistent before, during, and after pregnancy

Our primary study objective was to identify the overall changes to the T cell response using samples collected before, during, and after pregnancy among LTBI+ women. We first identified the frequency of CD4+ and CD8+ T cells from samples collected before, during, and after pregnancy (gating shown in **Figure S1A**). The frequency of CD4+ and CD8+ T cells was similar over these time points (**Figure 1B-C**). In WLHIV, the frequency of CD4+ T cells was lower than in women without HIV, irrespective of pregnancy status (**Figure S1B**) and CD8+ T cell frequency was increased (**Figure S1C**). COMPASS FS and PFS of CD4+ T cells from all samples strongly correlated with our LTBI measurements (**Figure 1E-F**).

### Mtb-specific CD4+ T cells polyfunctional responses decreased during the third trimester of pregnancy

Mtb induces a polyfunctional T cell response that may act independently of IFNγ, and these cells are critical for Mtb control (33). In Mtb-specific CD4+ T cells, PFS was significantly different before, during, and after pregnancy (**Figure 2A**, gating in Figure S2; p = 0.0084, Kruskal-Wallis test). PFS was significantly decreased in the third trimester compared with pre-pregnancy (p = 0.0005), second trimester (p = 0.0034) and post-pregnancy (p < 0.017), but not during the first trimester (p = 0.099; **Figure 2A**). FS results were similar to PFS (**Figure 2B**). We detected fewer Mtb-specific cytokine-producing subsets during third trimester, and very few subsets producing multiple cytokines simultaneously (COMPASS heatmaps shown in **Figure S3A**). We noted diminished single cytokine responses during the third trimester in IL-2+CD4+ cells compared with the second trimester (**Figure S3B**; p = 0.026) which trended lower compared with pre-pregnancy (p = 0.13), first trimester (p = 0.12) and post-pregnancy (p = 0.09). CD4+TNF+ or CD4+IFNγ+ responses were similar across pregnancy (**Figure S3C-D**). Therefore, using COMPASS, we were able to detect changes to Mtb-specific T cell responses that may be clinically important but not immediately detectable by analyzing single cytokines separately.

**Figure 2.**
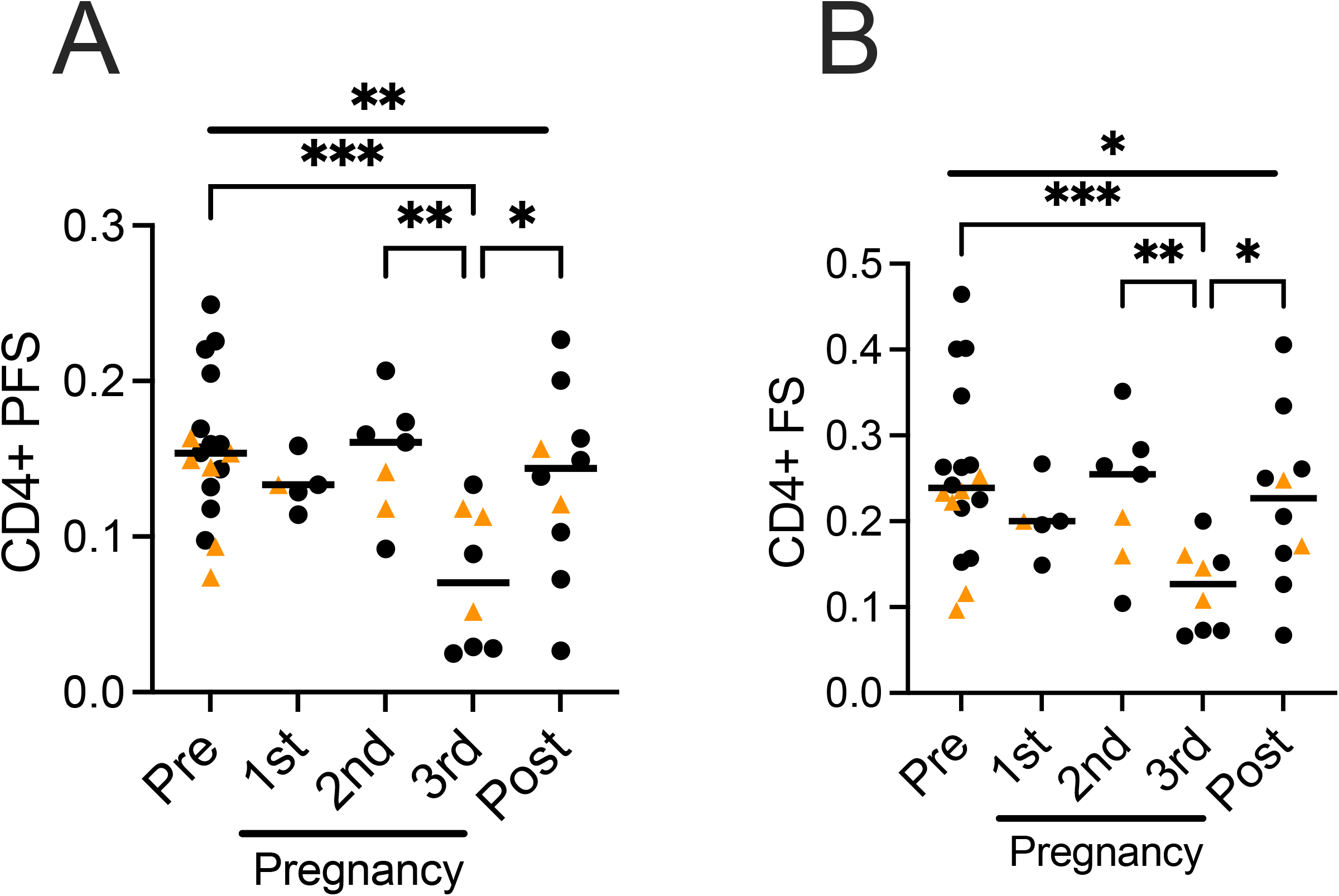
Mtb-specific CD4+ T cell responses are diminished during the third trimester of pregnancy. A) Polyfunctional scores (PFS) generated from COMPASS analysis, stratified by pregnancy trimester. *Black dots* indicate an HIV- study participant, while *gold triangles* indicate a WLHIV. Bars indicate median values. * P < 0.05, ** P < 0.01, *** P < 0.001, Kruskal-Wallis test. Column-to-column comparisons are made with Dunn’s test. B) Functional scores (FS) generated from COMPASS analysis, stratified by pregnancy trimester. *Black dots* indicate an HIV- study participant, while *gold triangles* indicate a WLHIV. Bars indicate median values. * P < 0.05, ** P < 0.01, *** P < 0.001, Kruskal-Wallis test. Column-to-column comparisons are made with Dunn’s test.

### CD4+ T cell responsiveness to a mitogen is altered during the third trimester

Clinical laboratory tests for LTBI include mitogen controls to assess overall T cell reactivity; impacts to mitogen induced CD4+ and CD8+ T cell responses during pregnancy directly alter LTBI diagnostic test performance (34). We compared T cell responses after stimulation with phorbol 12-myristate 13-acetate and ionomycin (PMA/ionomycin) over pregnancy in all study participants. We evaluated these T cell responses using traditional flow cytometry analysis, not COMPASS, as COMPASS is designed to detect antigen specific responses observed at low frequency. The frequency of IL2+CD4+ T cells responding to PMA/ionomycin were significantly changed over the duration of pregnancy (**Figure 3A**, p <0.0001, Kruskal-Wallis test). IL-2+CD4+ responses in the third trimester were increased compared to pre-pregnancy (p = 0.022), first trimester (p = 0.0006) and post-pregnancy (p < 0.0001). TNF+CD4+ responses were also significantly increased during the third trimester compared to first trimester (p = 0.0073) and post-pregnancy (p = 0.0002; **Figure 3B**, p = 0.0011 overall). IFNγ+CD4+ cells decreased across pregnancy (p = 0.0036). The third trimester response was significantly diminished compared to pre-pregnancy (p = 0.016), first trimester (p = 0.026), second trimester (p = 0.049), and post-pregnancy (**Figure 3C**; p = 0.0001). IL-2+, TNF+, and IFNγ+ CD4+ T cell frequencies were each significantly different before and after pregnancy (p = 0.006, p = 0.008, and p = 0.04, respectively), suggesting that systemic immune alterations of pregnancy begin during the first trimester and persist for weeks to months after birth. These data demonstrate distinct patterns of CD4+ T cell activation across pregnancy as compared to Mtb-specific T cells.

**Figure 3.**
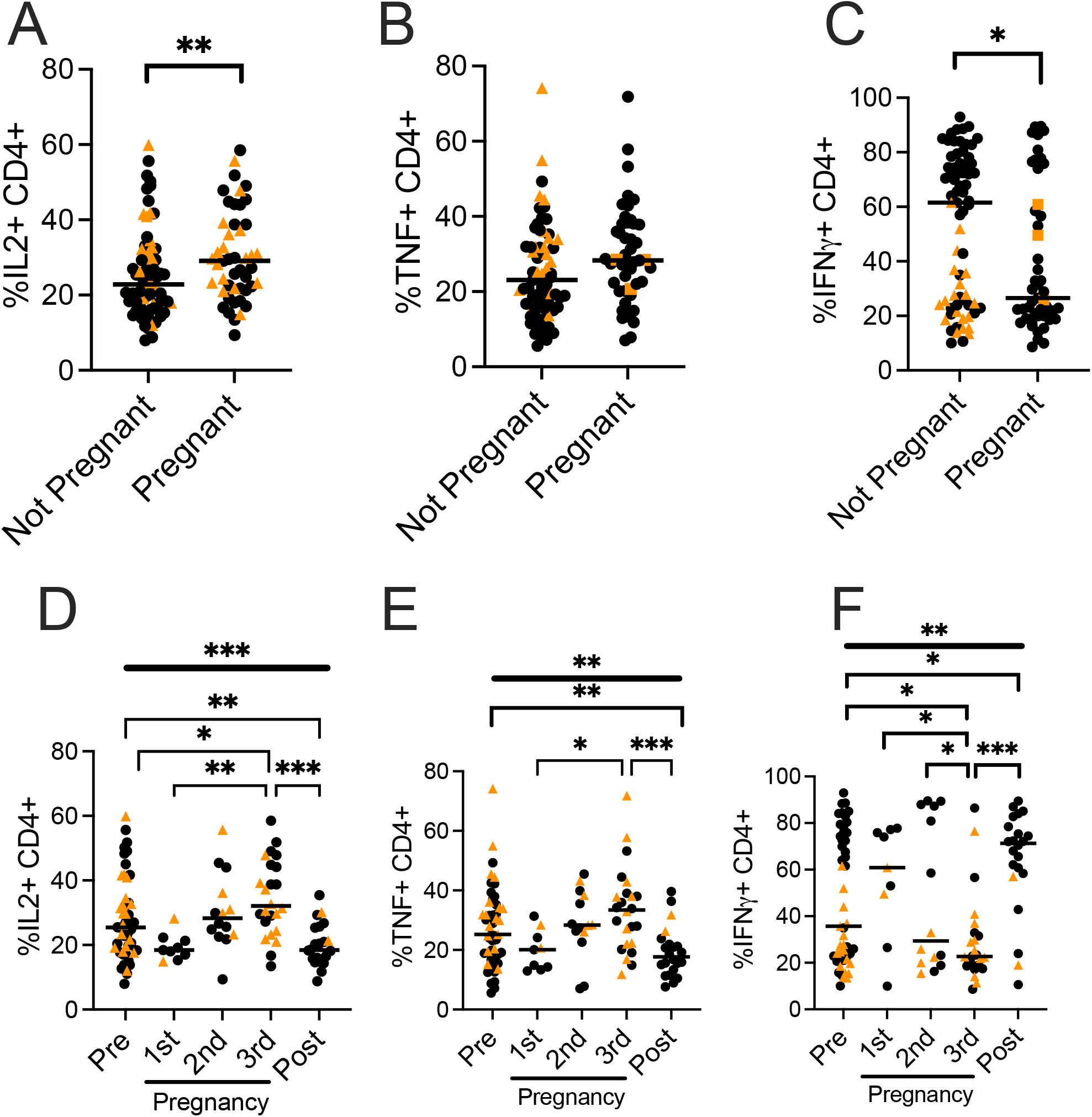
PMA/ionomycin-stimulated CD4+ T cell cytokine responses are dynamic across pregnancy. A-C) Proportion of A) IL-2+CD4+, B) TNF+CD4+, or C) IFNγ+CD4+ T cells, stratified by pregnancy status. *Black dots* indicate an HIV- study participant, while *gold triangles* indicate a WLHIV. Bars indicate median values. * P < 0.05, ** P < 0.01, *** P < 0.001, GEE with estimating equation, connected by *overhead bar*. D-F) Proportion of D) IL-2+CD4+, E) TNF+CD4+, or F) IFNγ+CD4+ T cells, stratified by pregnancy trimester. *Black dots* indicate an HIV- study participant, while *gold triangles* indicate an HIV+ study participant. Bars indicate median values. * P < 0.05, ** P < 0.01, *** P < 0.001, Overall significant by Kruskal-Wallis test demonstrated with top, *thick line without overhangs*. Column-to-column comparisons are made with Dunn’s test and shown with *thinner bars with overhangs*.

### Mtb-specific CD8+ T cells increase in activity during the second and third trimester of pregnancy

CD8+ T cells contribute to Mtb host defense and are measured in LTBI diagnostic tests (35, 36), but little is known about how pregnancy impacts Mtb-specific CD8+ T cell responses. We found low but detectable Mtb-specific CD8+ T cell cytokine responses in most samples from LTBI+ participants (**Figure S2**, gating strategy). The PFS and FS in CD8+ T cells changed significantly over the course of pregnancy (**Figure 4A-B**; p = 0.0033 and p = 0.038 respectively, Kruskal-Wallis test). Median PFS in CD8+ T cells was nearly undetectable before pregnancy and significantly increased in the second trimester compared with pre-pregnancy (p = 0.015), the first trimester (p = 0.023), and post-pregnancy (p = 0.034). A similar trend was observed in FS. The complexity of the CD8+ T cell response increased during the second and third trimester, and we detected subsets producing multiple cytokines simultaneously (**Figure S4A**). We were unable to detect shifts in IL-2, TNF, and IFNγ+ CD8+ T cell responses when evaluated singly, but we found a trend toward increased responses in each cytokine during the second and third trimester (**Figure S4B-D**). Taken together, Mtb-specific CD8+ T cell responses increased during the second and third trimester, while Mtb-specific CD4+ T cell responses diminished during the same period.

**Figure 4.**
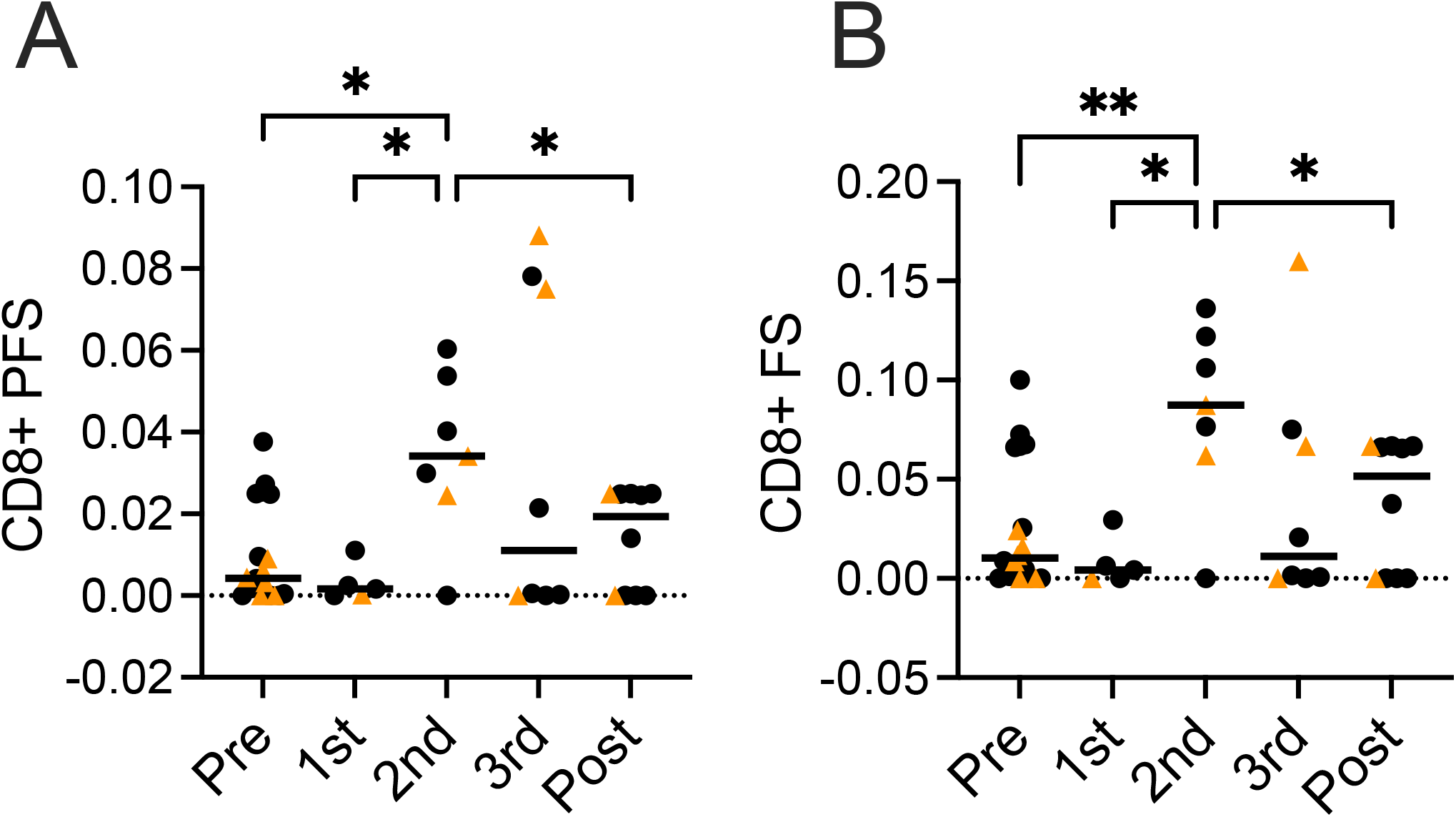
Mtb-specific CD8+ T cell polyfunctionality increases during the second trimester of pregnancy. A) Polyfunctional scores (PFS) generated from COMPASS analysis, stratified by pregnancy trimester. *Black dots* indicate an HIV- study participant, while *gold triangles* indicate a WLHIV. Bars indicate median values. * P < 0.05, ** P < 0.01, *** P < 0.001, Kruskal-Wallis test. Column-to-column comparisons are made with Dunn’s test. *Black dots* indicate an HIV- study participant, while *gold triangles* indicate a sample from WLHIV study participant. B) Functional scores (FS) generated from COMPASS analysis, stratified by pregnancy trimester. *Black dots* indicate an HIV- study participant, while *gold triangles* indicate a WLHIV. Bars indicate median values. * P < 0.05, ** P < 0.01, *** P < 0.001, Kruskal-Wallis test. Column-to-column comparisons are made with Dunn’s test.

### Mitogen-induced TNF+ CD8+ T cell responses increase during pregnancy

We measured the changes in PMA/ionomycin-stimulated CD8+ T cell responses over the course of pregnancy in all participants. IL-2+CD8+ responses were unchanged by pregnancy trimester (**Figure 5D;** p = 0.14, Kruskal-Wallis test). TNF+CD8+ T cell frequency changed over the course of pregnancy (**Figure 5E**; p = 0.0084) and increased in the third trimester compared with the first trimester (p = 0.015) and post-pregnancy (p = 0.0018). IFNγ+CD8+ T cell frequencies were similar across pregnancy (**Figure 5F**). These responses were distinct from the changes observed in Mtb-specific CD8+ T cells, where multiple subsets increased in frequency during the second and third trimester.

**Figure 5.**
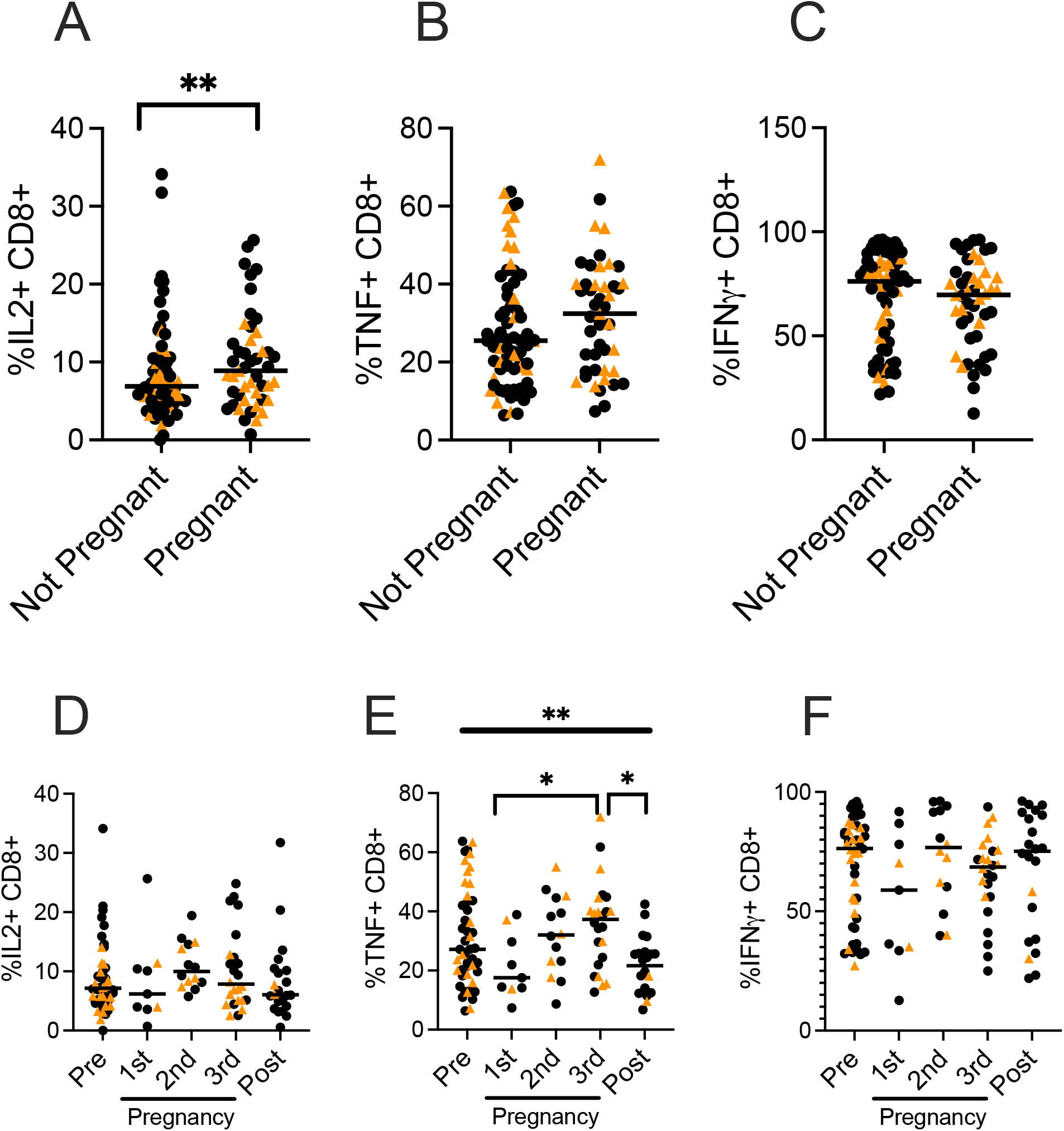
Cytokine responses in PMA/ionomycin-stimulated CD8+ T cells are stable during pregnancy. A-C) Proportion of A) IL-2+CD8+, B) TNF+CD8+, or C) IFNγ+CD8+ T cells, stratified by pregnancy status. *Black dots* indicate an HIV- study participant, while *gold triangles* indicate an HIV+ study participant. Bars indicate median values. * P < 0.05, ** P < 0.01, *** P < 0.001, GEE with estimating equation, connected by *overhead bar*. D-F) Proportion of D) IL-2+CD8+, E) TNF+CD8+, or F) IFNγ+CD8+ T cells, stratified by pregnancy trimester. *Black dots* indicate an HIV- study participant, while *gold triangles* indicate an HIV+ study participant. Bars indicate median values. * P < 0.05, ** P < 0.01, *** P < 0.001, Kruskal-Wallis test. Column-to-column comparisons are made with Dunn’s test.

### Mtb-specific effector memory T cell populations diminish during the third trimester

T cells display cell surface markers CD45RA and CCR7 as markers of T cell memory phenotype (37). Naïve (CCR7+CD45RA+), central memory (Tcm; CCR7+CD45RA-), and effector memory (Tem; CCR7-CD45RA-) T cells display distinct functional profiles and tissue homing sites, and Tcm are important for Mtb control (38-40). Naïve and Tcm produce little IFNγ, but Tem does not provide extended protection against disease. The frequency of total CD4+ and CD8+ T cells expressing CD45RA and CCR7 did not change over pregnancy (**Figure S5A-C**). We evaluated CD45RA and CCR7 expression in Mtb-specific CD4+ T cells, which were ESAT-6 and CFP-10 peptide incubated, cytokine-producing, CD4+ T cells from LTBI+ individuals. The proportion of Mtb-specific naïve and Tcm CD4+ T cells remained stable (**Figure 6A-B**), but we observed reduced Tem frequency during the third trimester (**Figure 6C**, p = 0.037; Kruskal-Wallis test) compared to pre-pregnancy (p = 0.049), first trimester (p = 0.005), second trimester (p = 0.014), and postpartum (p = 0.031). Viewed proportionally, naïve CD4+ cells increased while Tem decreased during the third trimester of pregnancy compared to the first trimester, second trimester, and postpartum, without substantial changes to Tcm frequency (**Figure 6D)**. We did not detect adequate Mtb-specific CD8+ T cells to evaluate T cell phenotypes in this population. In sum, Tem and IFNγ+CD4+ cells were reduced during the third trimester, but without increases in the Tcm population, which is important for Mtb host defense and may predict worsened Mtb control.

**Figure 6.**
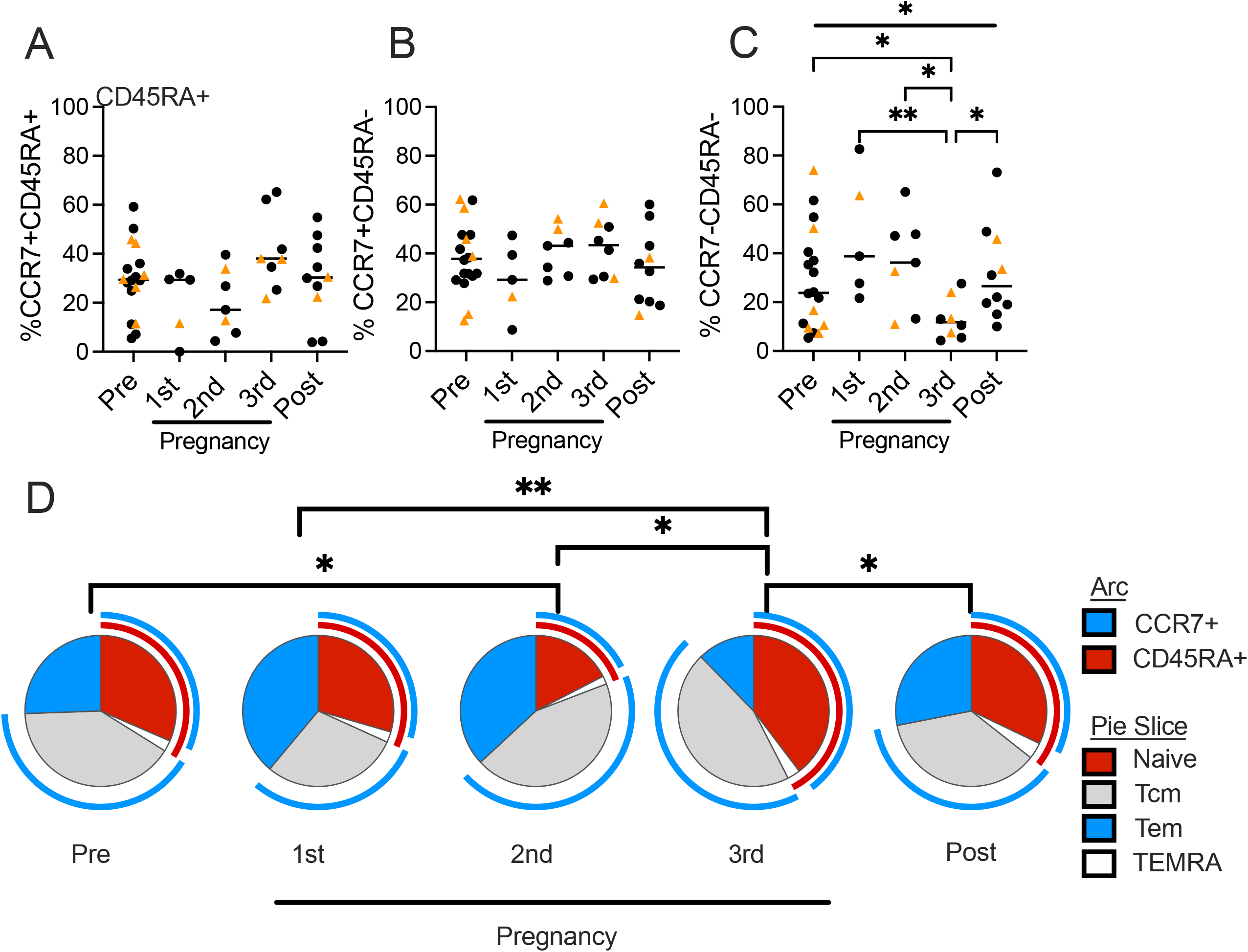
Proportions of Mtb-specific naïve CD4+ T cells increase and effector memory T cells decrease during the third trimester of pregnancy. A-C) Overall proportion of A) naïve (CCR7+CD45RA+), B) central memory (T_CM_; CCR7+CD45RA-), and C) effector memory (T_EM_; CCR7-CD45RA-) among all cytokine-producing CD4+ T cells after re-stimulation with ESAT-6 and CFP-10 antigens, stratified by pregnancy status. *Black dots* indicate an HIV- study participant, while *gold triangles* indicate a sample from WLHIV study participant. * P < 0.05, ** P < 0.01, *** P < 0.001, Kruskal-Wallis test. Column-to-column comparisons are made with Dunn’s test. D) Pie chart of Boolean-gated T helper memory phenotype frequencies, comparing the third trimester with other timepoints, connected by *overhead bar. Blue* arc indicates CCR7+ cells, *red* arc indicates CD45RA+ cells. *Red* pie slice: Naïve CD4+ T cells (Naïve); *blue* pie slice: effector memory CD4+ T cells (Tem); *gray* slice: central memory T cell (Tcm); *white* slice: TEMRA cell (TEMRA). * P < 0.05, ** P < 0.01, *** P < 0.001, permutation test of 10000 iterations.

### Nonspecific T cell activation increases during pregnancy

Increased nonspecific CD4+ T cell activation is a correlate of risk for developing Mtb disease (41, 42). We measured the frequency of HLA-DR+CD38+ CD4+ and CD8+ T cells across pregnancy, using “flow minus one” controls (**Figure S6A**). The frequency of HLA-DR+CD38+ CD4+ T cells significantly changed over pregnancy (**Figure 7A**; p < 0.0001, Kruskal-Wallis test), highest in the third trimester compared to pre-pregnancy (p = 0.0068), first trimester (p < 0.0001), second trimester (p = 0.007), or postpartum (**Figure 7A**; p < 0.0001). HLA-DR+CD38+ CD8+ T cell frequency also significantly changed similarly to CD4+ T cells (**Figure 7B**; p < 0.0001, Kruskal-Wallis test). In both CD4+ and CD8+ T cells, activation markers decreased from pre-pregnancy to the first trimester, and WLHIV demonstrated the greatest T cell activation throughout pregnancy. Therefore, T cell activation decreases at the outset of pregnancy, then increases over time until delivery.

**Figure 7.**
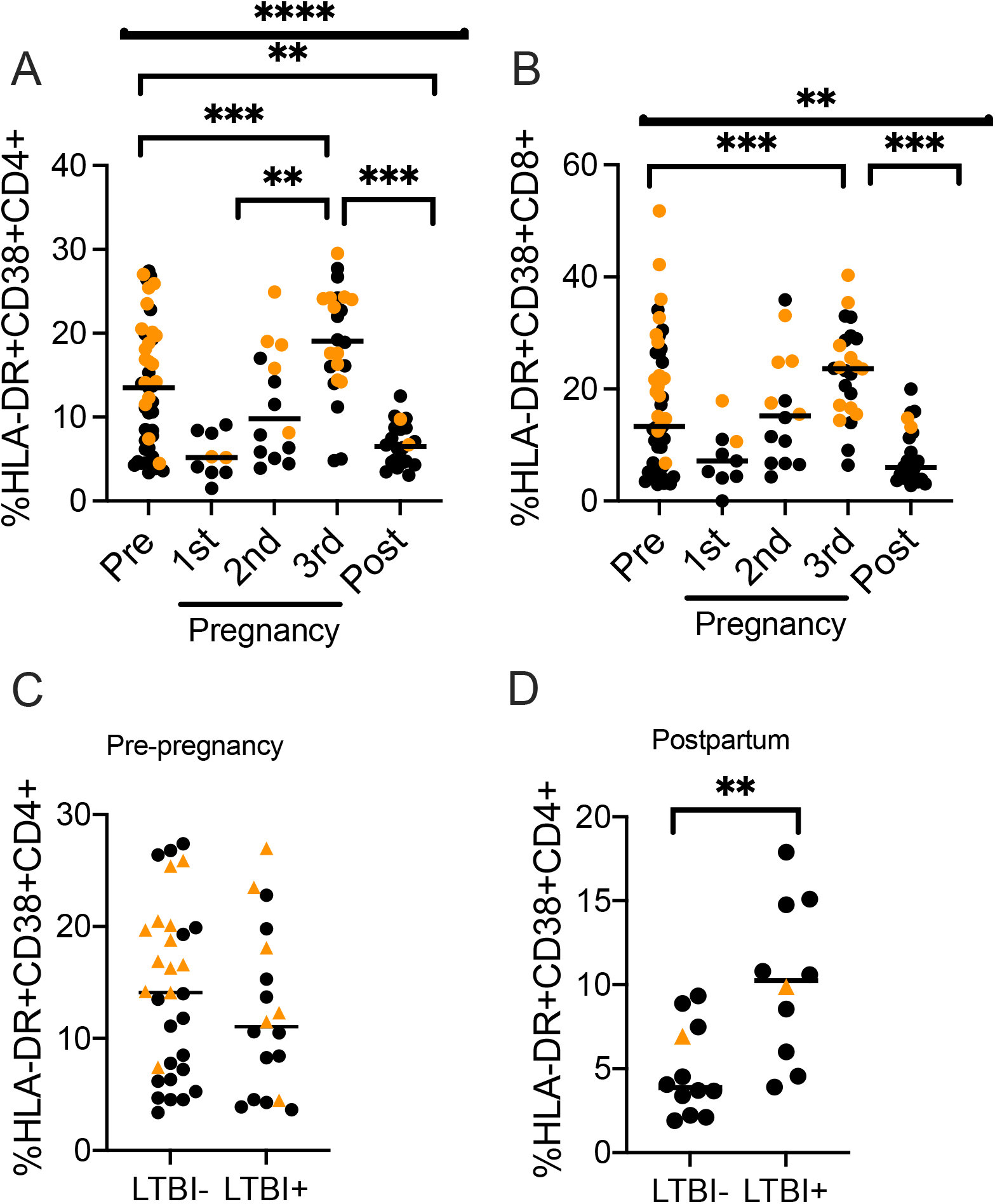
Nonspecific T cell activation increases during pregnancy and LTBI+ women demonstrate increased T cell activation postpartum. A) Proportion of HLA-DR+CD38+ CD4+ T cells, stratified by pregnancy status. B) Proportion of HLA-DR+CD38+ CD8+ T cells, stratified by pregnancy status. C) Proportion of HLA-DR+CD38+ CD4+ T cells from samples collected pre-pregnancy, stratified by LTBI status of the participant. D) Proportion of HLA-DR+CD38+ CD4+ T cells from samples collected postpartum, stratified by LTBI status of the participant. *Black dots* indicate an HIV- study participant, while *gold triangles* indicate sample from WLHIV. Thick bars correspond to overall changes (Krukal-Wallis test). *Bars with overhangs* indicate analysis of differences between columns (Dunn’s test). Bars indicate median values. * P < 0.05, ** P < 0.01, *** P < 0.001, Kruskal-Wallis test. Column-to-column comparisons are made with Dunn’s test in multiple group comparisons or Mann-Whitney U-test for tests between two groups.

We leveraged the observation that CD4+ T cell activation is a biomarker for TB risk to evaluate if the immune changes of pregnancy might increase the risk for TB progression from asymptomatic to symptomatic disease. We hypothesized that selective increases in nonspecific T cell activation would be present in LTBI+ women after pregnancy due to their increased risk for postpartum Mtb progression but not for LTBI-women. Pre-pregnancy, the proportion of HLADR+CD38+CD4+ T cells was equivalent between LTBI- and LTBI+ individuals (**Figure 7C**). Postpartum, LTBI+ individuals demonstrated significantly higher HLA-DR+CD38+ CD4+ T cells as compared to LTBI-individuals (**Figure 7D**; p = 0.0026, Mann-Whitney test). Immune changes of pregnancy in LTBI+ individuals may substantially contribute to overall T cell activation in the postpartum period and predict increased risk for TB progression.

## Discussion

In this study, we performed a comprehensive analysis of pregnancy stage on Mtb-specific T cell phenotypes in sub-Saharan African women including women living with HIV. These data are, to our knowledge, the first longitudinal assessment of systemic T cell responses to Mtb in pregnancy that includes samples collected prior to pregnancy. Taken in sum, we found that late pregnancy is associated with multiple changes to Mtb-specific T cell function: 1) overall Mtb-specific CD4+ T cell responses diminished and simplified; 2) Mtb-specific CD4+ Tem, which are cells that strongly produce IFNγ, decreased (37); 3) Mtb-specific CD8+ T cell responses increased, and 4) nonspecific T cell activation increased. (**Figure 8**). Nonspecific T cell activation increased in LTBI+ individuals compared to LTBI-individuals postpartum, despite being equivalent pre-pregnancy. Increased nonspecific T cell activation, a correlate of risk for Mtb disease in LTBI+ individuals, supports the observation that TB risk increases in the early postpartum period (41). Together, late pregnancy induces complex changes in the known T cell response to Mtb that have been associated with TB progression in other populations, and we found that LTBI+ women develop increased correlates of risk for TB in the immediate postpartum period.

**Figure 8.**
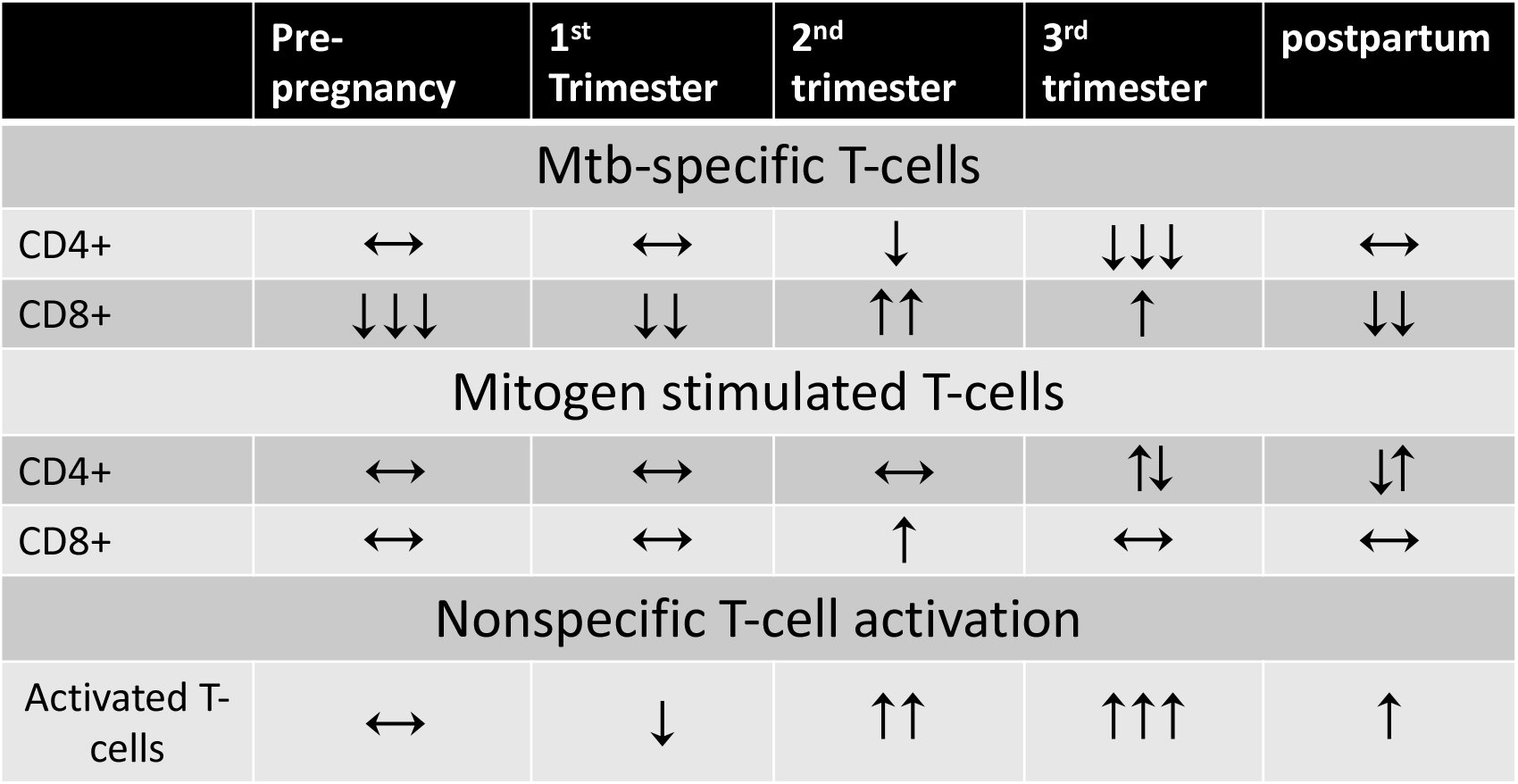
Summary of overall Mtb-specific T cell responses during differing stages of pregnancy. We observed distinct changes in Mtb-specific T cell responses as compared to mitogen-stimulated T cells. Arrows indicate the overall frequency of the responses measured. A downward arrow followed by an upward arrow indicates diminished IL-2/TNF responses with simultaneous increases in IFNγ+ cell frequency, and an upward arrow followed by a downward arrow indicates increases in IL-2/TNF+ T cell frequency with decreases in IFNγ+ responses.

We observed distinct changes between Mtb-specific and more generalized mitogen (PMA/ionomycin)-induced T cell cytokine responses. These results have direct implications toward the performance of LTBI diagnostics in pregnant women. Mtb-specific CD4+ T cell responses decreased during the third trimester but increased in the second trimester, and Mtb-specific CD8+ T cell responses were low but increased during the second and third trimester. By contrast, overall mitogen responses in CD4+ T cells, especially IL-2 responses, increased during late pregnancy. In other cohorts, discordance between the two commonest LTBI tests, TST and IGRA, has been noted especially among women in the late second and early third trimester, which may be partly due to the changes in the immune responses of pregnancy (34, 43). Part of the difference may be due to the absence of mitogen testing in most TSTs. Understanding this discordance in pregnancy may have implications for TB progression. Among those with positive IGRA but negative TST tests, Mtb-specific IL-2 responses associated with postpartum TB progression from asymptomatic infection to symptomatic disease (44). In combination, discordance between Mtb-specific and nonspecific immune responses may contribute to increased false-negative LTBI testing during the third trimester and may contribute to Mtb progression.

Conversely, we found that Mtb-specific CD8+ T cell responses increased during late pregnancy when CD4+ T cell responses were diminished. The role of CD8+ T cells in TB pathogenesis is uncertain. In children, Mtb-specific CD8+ T cell frequencies are correlated with Mtb disease (45). In animals, CD8+ T cell responses correlate strongly with Mtb bacterial burden in the lung (46, 47). CD8+ T cells recognize antigens presented on major histocompatibility complex I, which presents intracellular antigens. CD8+ T cells preferentially recognize antigen-presenting cells with an increased bacterial burden (48). During active intracellular infection, these responses may increase proportionally. It is unclear if some of these responses reflect emerging TB disease or if there are general postpartum factors that enhance CD8+ responses. These data suggest that CD8+ T cell responses may increase to compensate for diminished CD4+ T cell responses but may be unable to fully reproduce CD4+ T cell immune control.

The strengths of this study include its longitudinal nature, including samples obtained before, during, and after pregnancy. Most studies of the effects of pregnancy enroll women during pregnancy and often focus on the second and third trimester, which do not capture pre-pregnancy and first trimester periods. Several T cell phenotypes change significantly between pre-pregnancy and the first trimester, and other phenotypes differed between pre-pregnancy and postpartum. Local inflammation is critical for embryonic implantation, which suggests that both local and systemic immune changes begin immediately and are prolonged post-partum (49, 50). Another strength of this study was the use of COMPASS. COMPASS analysis revealed two findings that were not observable using standard data analysis techniques. This sensitive analysis permitted us to observe the diminished complexity of the Mtb-specific CD4+ T cell response during the third trimester and increases in overall CD8+ activity. Taken together, no single cytokine response accounted for the changes to the Mtb-specific T cell response, suggesting broad changes in the functional capacity of T cells responding to antigen during the third trimester.

Weaknesses of this study include the lack of TB disease outcomes data. This study used repository samples from a trial not originally designed to evaluate TB. However, we performed detailed immune characterization of the changes to the Mtb-specific immune response across pregnancy. Importantly, LTBI+ women had increased T cell activation postpartum, when compared with LTBI-women, which is a validated biomarker of TB disease progression in other cohorts (23). IGRA or TST was not performed in the parent study, which prevents correlation with clinically validated LTBI testing. However, the method we used in this study to identify LTBI+ participants is a well-accepted alternate approach that correlates with LTBI diagnosis (30, 31). Generally, flow-based testing is more sensitive to detect prior exposure to Mtb antigens than IGRA, so this approach permits us to evaluate a broader range of individual immune responses. Errors introduced by the inclusion of individuals without preexisting Mtb exposures are likely to reduce the sensitivity of this analysis to detect changes. We performed sensitivity analyses by raising the threshold for LTBI+ identification, but this did not substantially change the number of participants considered to have LTBI. Due to the limited sample size of this study, we did not perform formal analysis to investigate the impact of HIV on immune responses during pregnancy. Although HIV status was associated with reduced CD4+ T cell count and increased T cell activation, we found immune changes of pregnancy were generally independent of HIV status, though direct comparisons were limited due to small sample size.

TB is the third leading cause of death among women of child-bearing age in high burden areas and is associated with morbidity and mortality in pregnant women and their children, especially among those affected by HIV (1, 43). Our study provides evidence that systemic changes in the T cell response during the third trimester may contribute to increased risk for TB progression. T cell cytokine responses are reduced and simplified, T cell memory phenotypes skewed toward naïve phenotypes, and nonspecific activation is enhanced, which all are independently associated with TB risk. Recent data suggest that isoniazid preventative therapy is associated with adverse pregnancy outcomes, with a signal that this may occur more often if initiated early in pregnancy (51). As such, late pregnancy, which is associated with key immunologic changes for TB progression, may be an advantageous time to target for shorter course regimens. Understanding host factors that place pregnant mothers at risk for TB may provide insight into optimal public health strategies for TB control in this high-risk population.

## Supporting information

Supplemental menthod, figure legends and table 1

## Data Availability

Anonymized data from this study can be made available on request to the authors.

## Acknowledgements

The authors would like to thank the study participants and their families. We would like to acknowledge the support of Harald Haugen on database management of clinical samples. We would also like to acknowledge the clinical teams who obtained informed consent and collected samples for this biorepository. We would also like to thank Chetan Seshadri for helpful technical conversations.

