## Supplementary figures and images for "*Mycobacterium tuberculosis*-specific T cell responses are impaired during late pregnancy with elevated biomarkers of tuberculosis risk postpartum"

### Supplemental figure 1

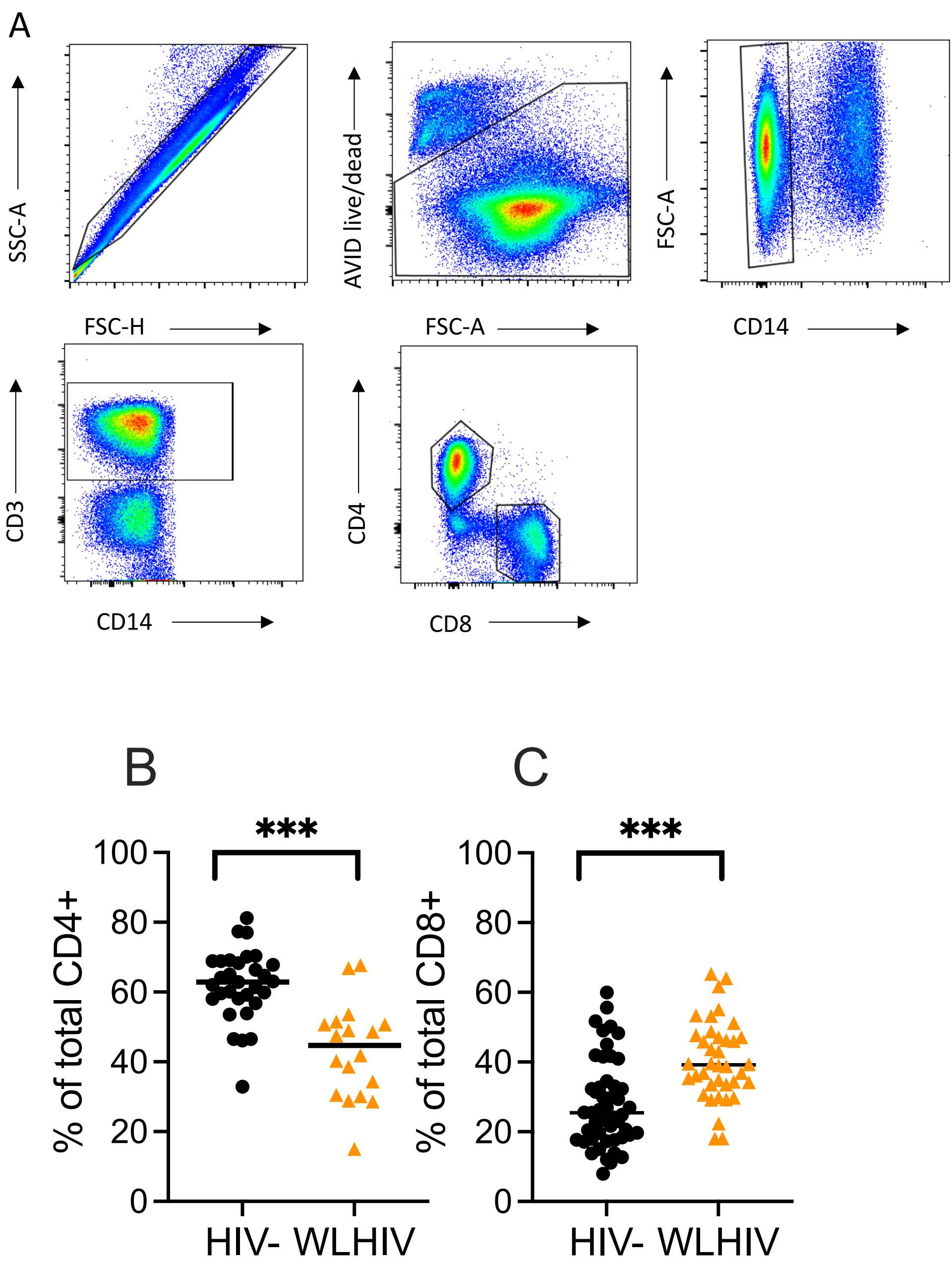

### Supplemental figure 2

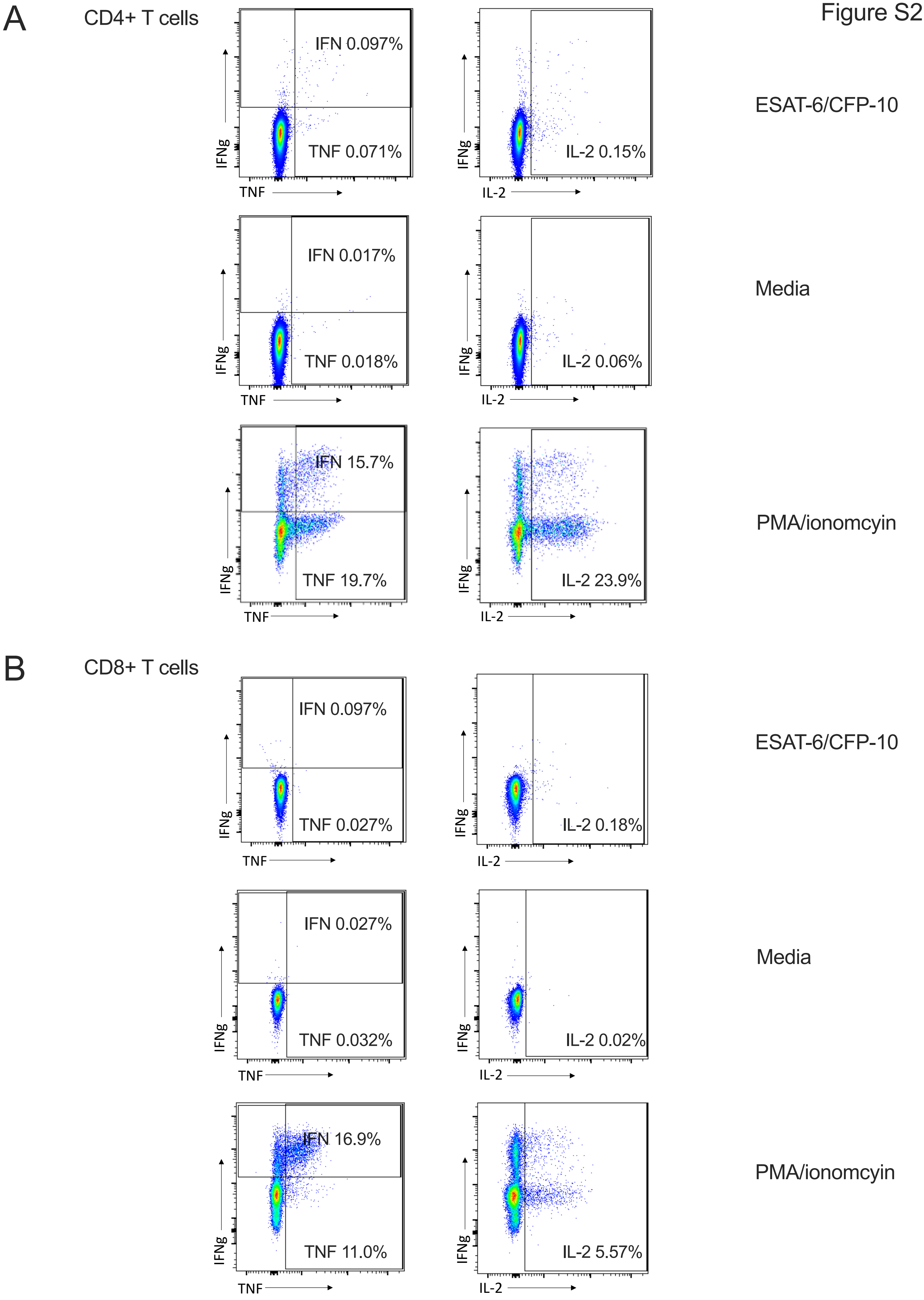

### Supplemental figure 3

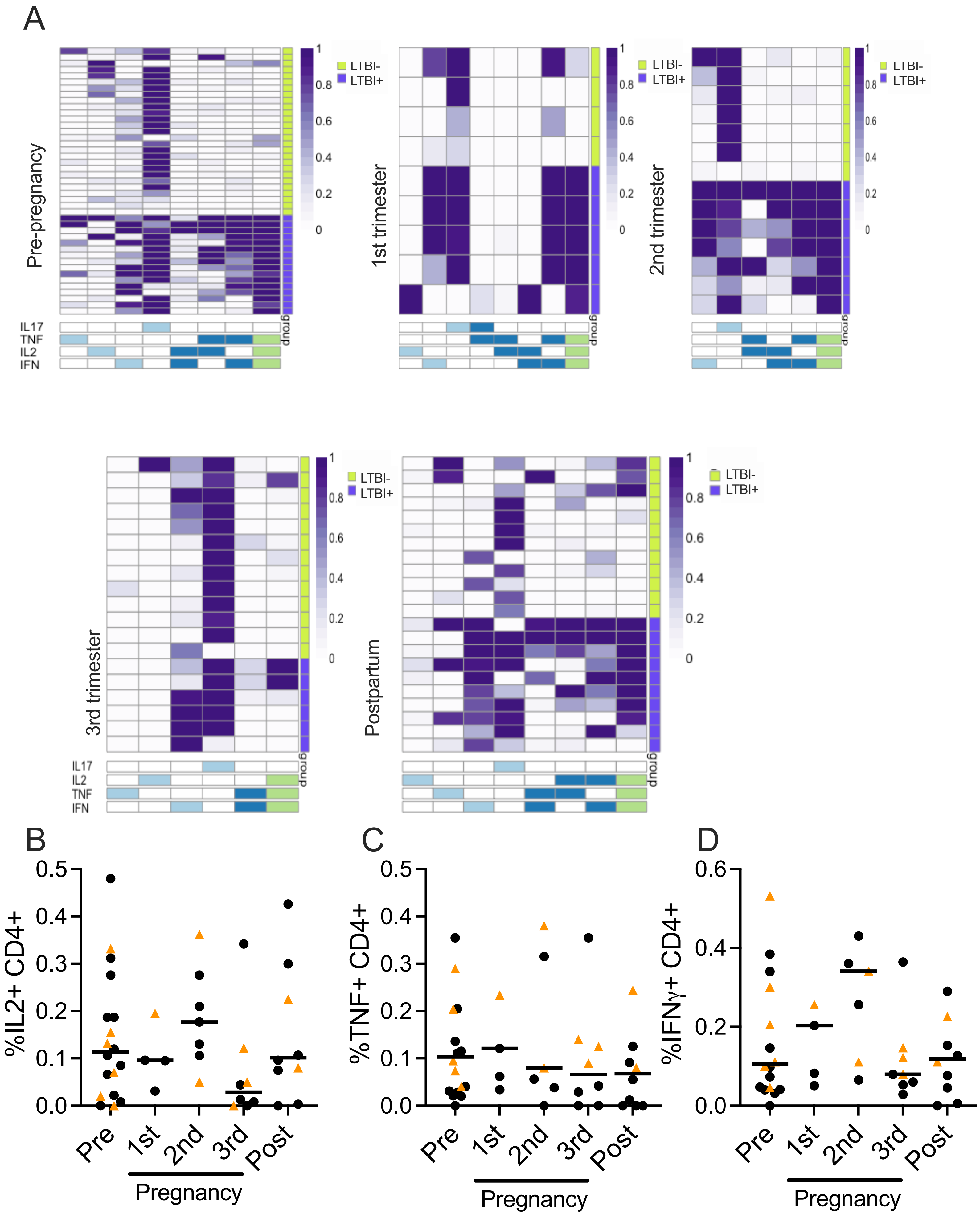

### Supplemental figure 4

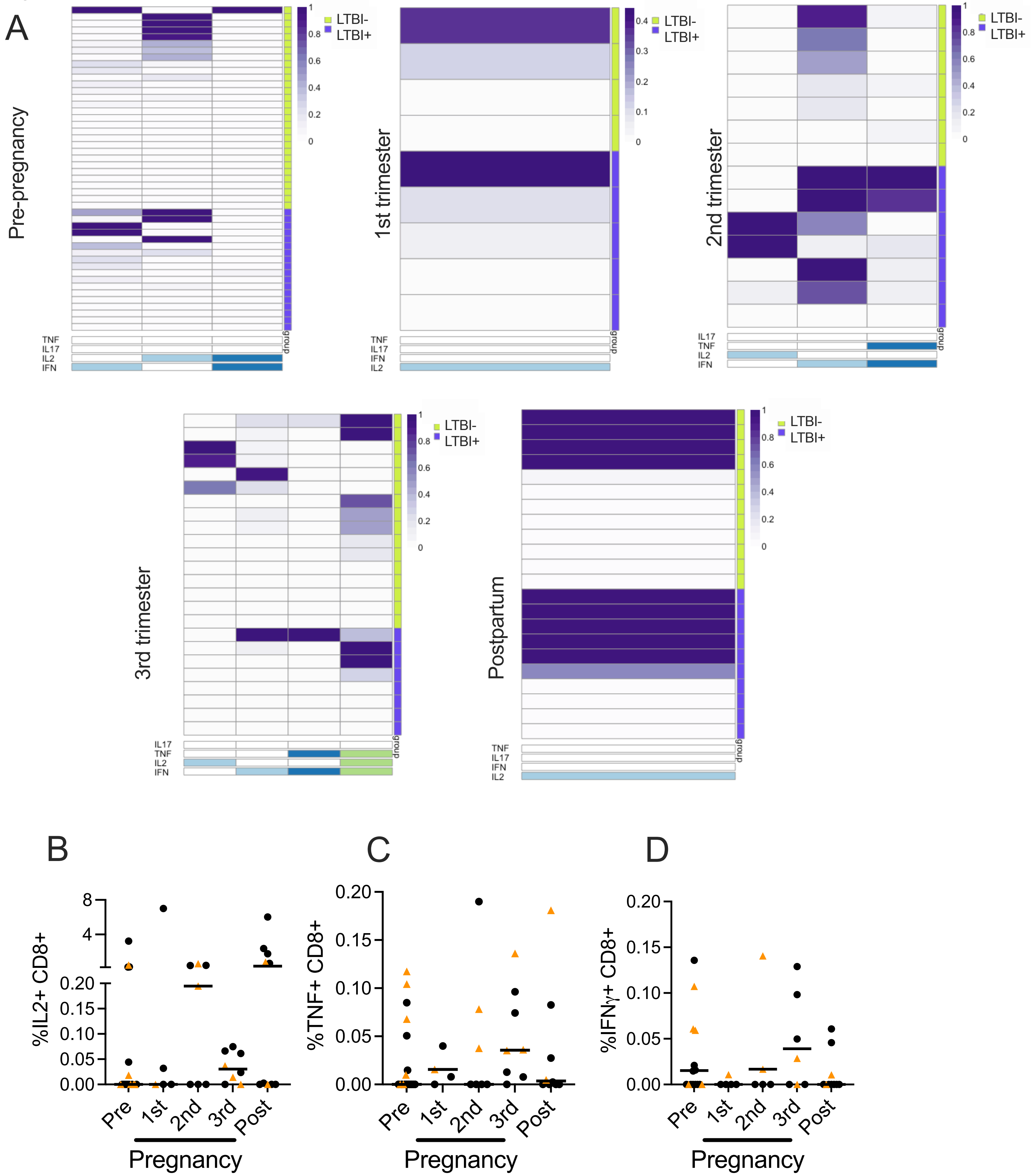

### Supplemental figure 5

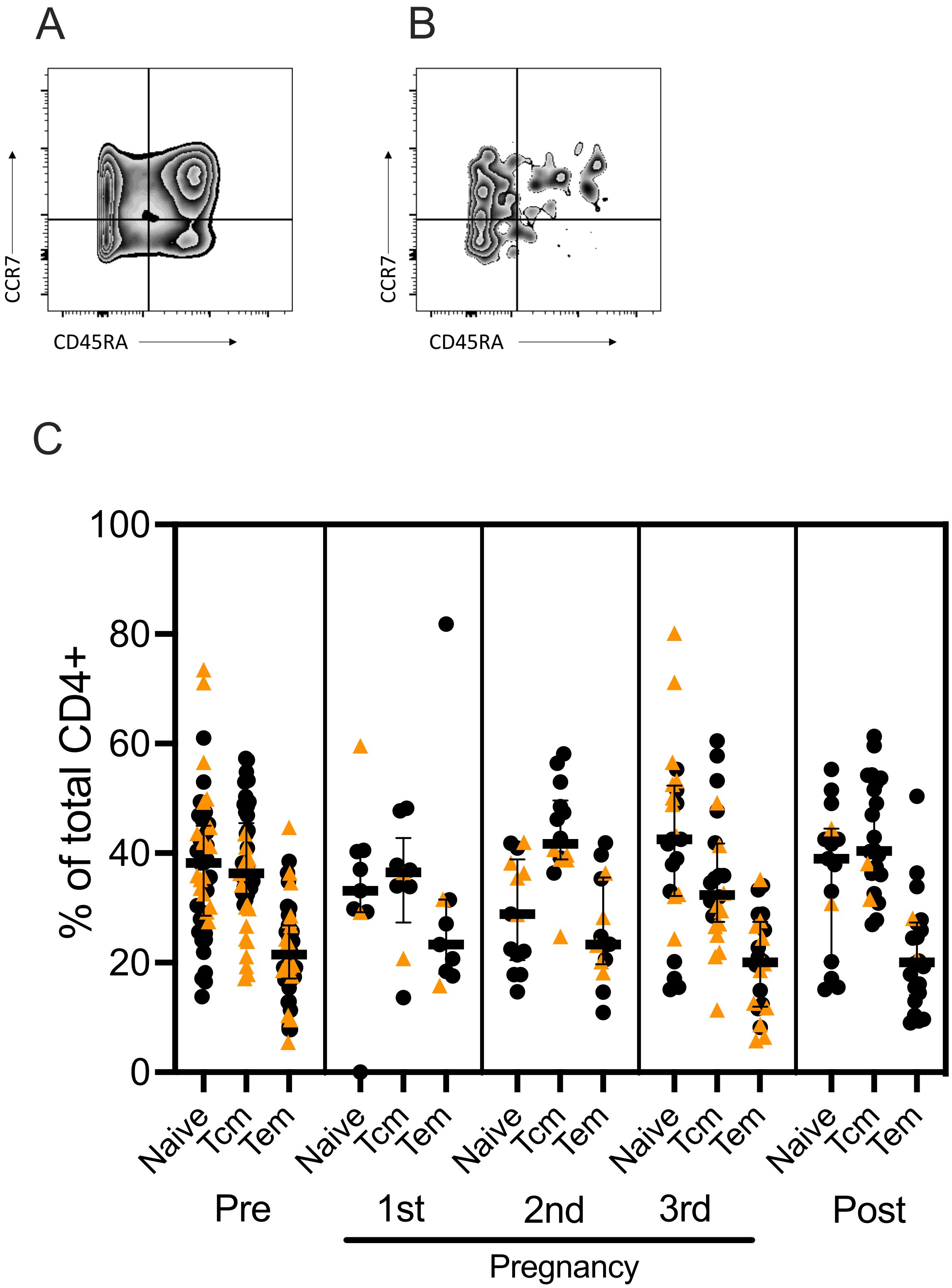

### Supplemental figure 6

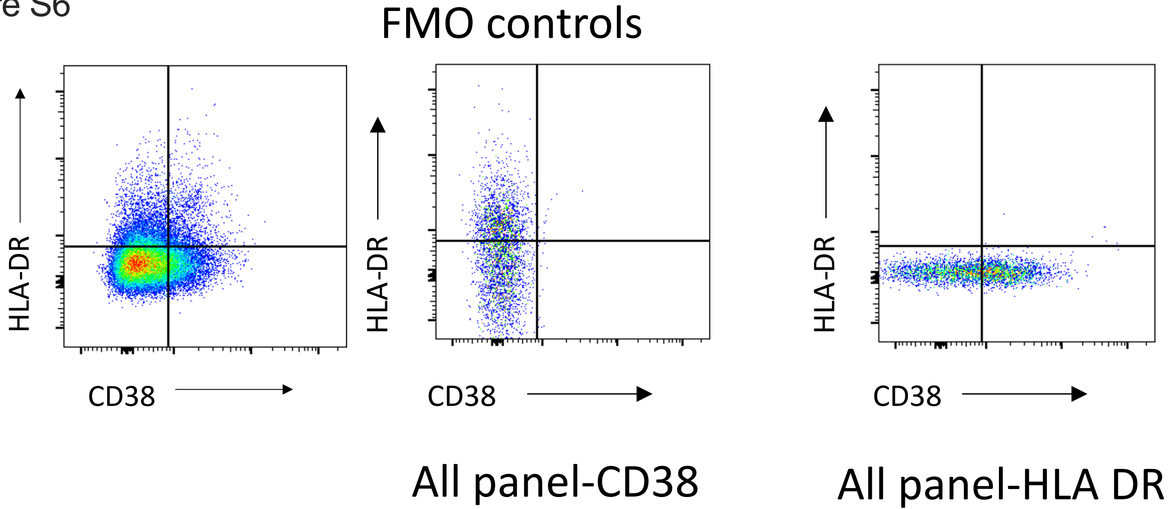
