## Supplemental menthod, figure legends and table 1 for "*Mycobacterium tuberculosis*-specific T cell responses are impaired during late pregnancy with elevated biomarkers of tuberculosis risk postpartum"

*Both authors contributed equally

**Supplemental Methods**

*Study Population*

The protocol for this study was approved by the University of Washington Human Subjects Review Committee and ethics review committee at each study site. All participants provided written informed consent in English or their native language.

The Partners PrEP Study was a randomized clinical trial of antiretroviral pre-exposure prophylaxis (PrEP) in HIV serodiscordant couples to reduce risk of HIV acquisition (28). A total of 4758 participants were enrolled from 9 sites in Kenya and Uganda between 2007 and 2012. HIV-infected partners with CD4 cell count ≥250, without history of AIDS defining diagnoses or current use of antiretroviral therapy (ART) were eligible. HIV-uninfected participants underwent monthly HIV testing. Whole blood was collected every six months at 5 of 9 study sites in Kenya and Uganda and processed for peripheral blood mononuclear cells (PBMC). Participants who were pregnant at enrollment were excluded from the parent trial, but those with incident pregnancy continued in follow-up. HIV-uninfected women had urine pregnancy testing at enrollment and monthly thereafter; women living with HIV (WLHIV) had urine collected quarterly. WLHIV not already on ART who became pregnant were referred for prevention of maternal to child transmission services. A total of 1495 women had incident pregnancies in this study. Participants were eligible for this analysis if they had paired samples available within 6 months prior to incident pregnancy, during, and within 6 months post pregnancy. For women with only 2 timepoints available, we prioritized those with pregnancy samples available at >20 weeks gestation, hypothesizing that the greatest immunologic changes of interest would occur during the later stages of pregnancy. (**Figure 1A**).

*Reagents*

RPMI 1640 medium, L-glutamate were obtained from Life Technologies. Fetal calf serum was obtained from Atlas Technologies. Whole cell lysate from *M. tuberculosis* strain H37Rv was obtained as part of National Institutes of Health, National Institute of Allergy and Infectious Diseases Contract No. HHSN266200400091C, entitled Tuberculosis Vaccine Testing and Research Materials (Colorado State University, Fort Collins, CO). Pooled peptides were obtained from BEI Resources, NIAID, NIH: Peptide Array, Mycobacterium tuberculosis CFP-10 Protein, NR-34825 and Peptide Array, Mycobacterium tuberculosis ESAT-6 Protein, NR-34824.” PMA and ionomycin was obtained from eBioscience. Antibodies (clones and source) for flow cytometry with intracellular cytokine staining are shown in **Table S1**.

*Flow Cytometry*

Cryopreserved PBMCs were thawed, washed and rested in Roswell Park Memorial Institute RPMI 1640 media containing 10% heat-inactivated fetal bovine serum (FBS) overnight prior to antigen stimulation at a conc. of 2 x 10^6^ cells /mL at 37°C in humidified incubator supplemented with 5% CO_2_. The following morning the PBMCs were counted with Guava easyCyte (Millipore) using Guava viacount reagent (Luminex) and guavasoft v.2.6 software. Samples with fewer than 66% viability were discarded. Partners cryopreserved PBMCs demonstrated average viability of 85-90%, and many studies have used these samples to successfully evaluate systemic and mucosal T cell responses to HIV-exposure as well as regulatory T cell activity in HIV-exposed uninfected individuals. Cells were plated at a minimum concentration of 1 x 10^6^ per well into 96-well U-bottom plate and stimulated with a pool of early secretory MTB antigen (ESAT-6) and culture filtrate protein (CFP-10) peptides in a 1:1 ratio (BEI resources), PMA (25ng/mL)/ionomycin(1ug/mL) was used as a positive control, or DMSO (0.5%) was used as a negative control. In addition, co-stimulatory antibody anti-CD28/49d, cytokine secretion inhibitor Brefeldin A and Monensin were added to each stimulation cocktail. Each stimulation was performed for 6h at 37 °C with 5% CO_2_ after which ethylenediamine tetra-acetic acid (EDTA), at a final concentration of 2 mM, was added to disaggregate cells. The cells were stored at 4°C overnight. The following day the cells were washed with PBS and then stained for 20 min at room temperature with Avid Live/Dead (Invitrogen), prepared according to manufacturer’s instruction. After that, cells were lysed and permeabilized with FACS Lyse and FACS Perm-II buffer respectively. Cell staining was performed as per previously optimized and validated panel at 4°C for 30 min except CCR7, for which the incubation temperature was 37°C (29). Cells were fixed in 1% paraformaldehyde and acquired on a BD LSRFortessa (BD Biosciences), equipped with a high-throughput sampler and configured with blue (488 nm), green (532 nm), red (628 nm), violet (405 nm) and ultraviolet (355 nm) lasers using standardized good clinical laboratory practice. Samples with same patient ID were measured concurrently to avoid batch effects.

*Statistical Analysis*

Flow cytometry data was analyzed in FlowJo™ v10.7.1 Each sample were compensated and gated manually. Data were then exported to Excel where cytokine positive samples were identified if frequency of IFNγ production at least doubled in the peptide (ESAT-6/CFP-10) stimulation group compared with negative control, a well-established, sensitive method for determining prior exposure to Mtb (30). Median values of T cell responses were compared between timepoints of interest using Kruskal-Wallis tests to determine overall statistical significance followed by Dunn’s test comparing individual columns as a secondary analysis to define mechanisms of action. Statistical analysis was performed in GraphPad Prism 8.1 or Stata 14.1. All *p*-values presented are two sided, and *p* < 0.05 was the cutoff for statistical significance unless otherwise stated in the text. Cytokine combinations were assessed using SPICE (simplified presentation of incredibly complex evaluations) software and data are reported after background subtraction. COMPASS, a Bayesian statistical method for evaluating overall antigen-specific T cell responses, was used to identify antigen-specific T cell subsets and create an overall score describing the magnitude of the T cell response as previously described (31, 32).

**Supplemental Information.**

**Supplemental Figure 1.** WLHIV demonstrate reduced proportions of total CD4+ T cells and increased CD8+ T cells. PBMC from pregnant women before, during, or after pregnancy were thawed, rested overnight, then stimulated with media, pooled, overlapping 15 amino acid long peptides encompassing the Mtb ESAT-6 and CFP-10 proteins, or PMA and ionomycin for 6 hours, then fixed, permeabilized, and stained with fluorescent antibodies.

A) The gating strategy is described. From *left* to *right*, singlets were selected, then live cells were selected. CD14+ cells were excluded and CD3+ cells were selected. CD4+ and CD8+ T cells were selected from this CD3+ population.

B-C) Proportion of total CD3+ T cells expressing A) CD4+ and B) CD8+ cell surface markers, stratified by HIV status. *** p < 0.001, Mann-Whitney U-test. Samples from HIV- individuals are labeled with *black circles* and samples from PLHIV are labeled with *orange triangles*.

**Supplemental Figure 2.** Representative dot plots of the frequencies of IL-2, TNF, and IFNγ responses after media, ESAT-6/CFP-10 pooled peptide+CD26/CD49d, or PMA/ionomycin stimulation from CD4+ and CD8+ T cells.

**Supplemental Figure 3.** Associated with Figure 2. A) COMPASS heatmaps of the probability of Mtb-specific responses in CD4+ T cells in each time point studied. Each row indicates a separate study volunteer, and each column represents a different Boolean combination of cytokines detected in this study. Purple shading indicated increased probability of a response in that individual. Participants with green bars on the right indicate that they were LTBI-, while those with lavender bars were LTBI+. This classification was made based upon detection of 2x increase in IFNγ+CD4+ T cells after ESAT-6/CFP-10 pooled peptide restimulation in timepoints outside of pregnancy, either pre-pregnancy or postpartum. Therefore, some participants classified as LTBI- may demonstrate IFNγ+ responses during pregnancy due to repeated measures over time.

B-D) Proportion of B) IL-2+CD4+, C) TNF+CD4+, or D) IFNγ+CD4+ T cells, stratified by pregnancy trimester. *Black dots* indicate an HIV- study participant, while *gold triangles* indicate an HIV+ study participant. Bars indicate median values. * P < 0.05, ** P < 0.01, *** P < 0.001, Kruskal-Wallis test. Column-to-column comparisons are made with Dunn’s test.

**Supplemental Figure 4.** Associated with Figure 4. COMPASS heatmaps of the probability of Mtb-specific responses in CD8+ T cells in each time point studied. Each row indicates a separate study volunteer, and each column represents a different Boolean combination of cytokines detected in this study. Purple shading indicated increased probability of a response in that individual. Participants with green bars on the right indicate that they were LTBI-, while those with lavender bars were LTBI+. This classification was made based upon detection of 2x increase in IFNγ+CD4+ T cells after ESAT-6/CFP-10 pooled peptide restimulation in timepoints outside of pregnancy, either pre-pregnancy or postpartum. Therefore, some participants classified as LTBI- may demonstrate IFNγ+ responses during pregnancy due to repeated measures over time.

B-D) Proportion of B) IL-2+CD8+, C) TNF+CD8+, or D) IFNγ+CD8+ T cells, stratified by pregnancy trimester. *Black dots* indicate an HIV- study participant, while *gold triangles* indicate an HIV+ study participant. Bars indicate median values. * P < 0.05, ** P < 0.01, *** P < 0.001, Kruskal-Wallis test. Column-to-column comparisons are made with Dunn’s test.

**Supplemental Figure 5.** Associated with Figure 6**.**

A) Representative figure for memory T cell surface marker expression, demonstrating CCR7 and CD45RA expression in total CD4+ cells without restimulation.

B) Representative Figure for gating in Figure 6C, demonstrating CCR7 and CD45RA expression in all CD4+ T cells expressing either IL-2, TNF, or IFNγ after 6 hours of restimulation with pooled peptides covering ESAT-6 and CFP-10 antigens.

C) Expression of CCR7 and CD45RA on total CD4+ T cells, stratified by pregnancy status.

**Supplemental Figure 6.** Associated with Figure 7. Representative gating for HLA-DR and CD38, with demonstration of flow minus one (FMO) controls, where full panels were stained with the exception of the fluorophore of interest. Representative images of the full staining panel, FMO of CD38, and FMO of HLA-DR are shown.

#### **Supplemental Table 1.** Key Resources.

| **Function** | **Antibody** | **Fluorophore** | **Manufacturer** | **Cat#** | **Clone** |
| --- | --- | --- | --- | --- | --- |
| Cell viability | AViD | AmCyan | Thermo Fisher | L34976 |  |
| T-cell lineage | CD3 | ECD | Beckman Coulter | IM2705U | UCHT-1 |
|  | CD4 | AF700 | eBioscience | 56-0049-42 | RPA-T4 |
|  | CD8 | PerCP Cy 5.5 | BD | 341051 | SK1 |
| Intracellular cytokines | IL2 | PE | BD | 559334 | MQ1-17H12 |
|  | IFNγ | APC | BD | 554702 | B27 |
|  | TNF | FITC | Beckman Coulter | 554512 | Mab11 |
| Monocyte | CD14 | BUV395 | BD OptiBuild | 740286 | M5E2 |
| T-cell memory | CCR7 | BV711 | BD | 563921 | 150503 |
|  | CD45RA | PE Cy7 | Biolegend | NC0215296 | HI100 |
| T-cell activation | HLA DR | BV605 | BD | 562845 | G46-6 |
|  | CD38 | BV785 | Biolegend | 303530 | HIT2 |
